# Extracting and calibrating evidence of variant pathogenicity from population biobank data

**DOI:** 10.1101/2024.08.14.24311911

**Authors:** Vineel Bhat, Tian Yu, Lara Brown, Vikas Pejaver, Matthew Lebo, Steven Harrison, Christopher A. Cassa

## Abstract

Genomic medicine requires a robust evidence base of variant phenotypic impacts, which remains incomplete even in extensively studied monogenic disease genes. Here, we evaluated the broad potential of using population cohort data to identify evidence that can be used in variant assessment. Across 41 genes related to 18 clinically actionable monogenic phenotypes, we calculated variant-level odds ratios of disease enrichment using data from 469,803 UK Biobank participants. We found significant differences in odds ratio values between ClinVar-labeled pathogenic and benign variants in 11 phenotypes, spanning both common and rare disorders. To facilitate clinical translation, we calibrated the strength of evidence provided by variant-level odds ratios to align with American College of Medical Genetics and Genomics (ACMG/AMP) interpretation guidelines (PS4 criterion), and found that odds ratios may reach ‘moderate’, ‘strong’, or ‘very strong’ evidence, varying by phenotype and gene. Overall, we found that 2.6% (N = 12,350) of participants harbor a rare VUS with at least ‘moderate’ evidence of pathogenicity – an indication of potentially unrecognized disease risk. Finally, by incorporating computational and functional data alongside population-based odds ratios, we identified variants that met criteria for clinical reclassification. Notably, using this approach, we identified that 12.4% of rare VUS in *LDLR* seen in participants meet diagnostic criteria to be classified as likely pathogenic, demonstrating its potential to scale the reclassification of VUS.

**Highlights:**

- We identify rare coding variants that alter risk across 41 genes related to 18 actionable phenotypes.
- We find enrichments of cases in variants of uncertain significance related to rare and common disorders.
- We calibrate the strength of biobank population evidence for use in sequence variant interpretation.
- We combine evidence to identify uncertain variants that can be reclassified as likely pathogenic.

## Introduction

Advancing genomic medicine relies on our ability to identify variants with sufficient clinical certainty that they can be labeled ‘pathogenic’ or ‘benign’.^1^ Diagnostic testing has identified many pathogenic variants which increase disease risk, providing valuable insight into the molecular basis of inherited disorders.^2^ However, even in established disease genes like *BRCA1, LDLR*, and *MSH2*, most variants have insufficient evidence to reach a clinical classification, and are consequently labeled ‘Variants of Uncertain Significance’ (VUS).^3,4^ While many of these variants may substantially increase disease risk, current clinical guidelines discourage this prognostic information from being communicated to patients or providers who have no specific indication for testing when following current clinical guidelines.^5^ This translational gap collectively prevents many patients from benefiting from genomic medicine, including optimized surveillance and therapeutic options.^4^

To standardize the information used in variant assessment, the American College of Medical Genetics and Genomics and the Association for Molecular Pathology (ACMG/AMP) sequence variant interpretation (SVI) guidelines systematically weigh and combine the available evidence of pathogenicity or benignity for each variant. These guidelines define various forms of evidence (*e*.*g*., population, functional, computational, or contextual evidence) and assign strength levels to each type (*e*.*g*., ‘supporting’, ‘moderate’, ‘strong’, ‘very strong’), based on the clinical certainty provided by each form of evidence. Importantly, no single source of evidence alone is considered sufficient to classify a variant as pathogenic. The SVI guidelines define population evidence as ‘strong’ evidence of pathogenicity for a variant when it is significantly enriched in disease cases over controls (ACMG/AMP PS4 criterion).^6^

Given that damaging variants in disease genes tend to be rare, population evidence of pathogenicity has most often been derived by counting the number of distinct probands from clinical cases, or measuring the extent of co-segregation of variants with disease within families. Some disease-specific Variant Curation Expert Panels (ClinGen VCEPs) have defined specialized interpretation criteria for population evidence for specific genes and phenotypes. For example, different numbers of probands that have a specific disorder can correspond to different strengths of evidence for a phenotype (*e*.*g*., 2-5 distinct familial hypercholesterolemia cases provide ‘supporting’ evidence of variant pathogenicity in *LDLR*).^2,7^ When considering the population enrichment of cases at the variant level, most often, a variant with an odds ratio ≥ 5.0 and lower 95% confidence bound ≥ 1 is considered strong evidence of pathogenicity, though these criteria may also differ by disorder or even gene. Recent work has considered how to make use of population data in variant interpretation, and the extent to which population cohorts can provide evidence of variant pathogenicity, but this approach has not yet been measured or calibrated at the gene or phenotypic level, or across a broad set of phenotypes.^8,9^

Dramatic increases in biobank size now provide statistical power to detect a broader spectrum of variant effect sizes, particularly for disorders and phenotypes which are more common or which are widely measured in population cohorts.^10^ Notably, endophenotypic effect sizes have been shown to discriminate between variants which are known to be pathogenic or benign in monogenic susceptibility genes, generally with larger effects.^11^ However, this information has yet to be generalized across a broader set of dichotomous phenotypes or calibrated for use with existing clinical guidelines, which has limited its use in variant assessment.

Here, we draw on population case data at scale from 469,803 UK Biobank participants to identify variants which are enriched in 18 actionable disorders. Specifically, we model enrichment of disease using odds ratios for each phenotype based on case data at the variant level. We then align this evidence of pathogenicity within the ACMG/AMP SVI diagnostic framework by systematically calibrating the strength of evidence provided by variant odds ratios in each gene, enabling its use in clinical translation. Finally, we combine this information with well-calibrated computational and functional evidence to identify the scale of VUSs that could potentially be re-classified. By extracting and aligning population evidence in an automated manner, this framework represents a powerful step forward toward eliminating VUS.

## Results

### Population characteristics

To calculate variant-level odds ratios based on observed population outcomes, we draw on data from 469,803 participants in the UK Biobank with available whole exome sequencing data. Population-level summary statistics are provided in **Table 1**. After quality control, we observed 56,144 rare, nonsynonymous variants across 18 disorders and 41 genes, affecting 409,003 participants collectively.

**Table 1:** Population characteristics. Population characteristics for the UK Biobank cohort with available whole exome sequencing data that were considered in our analyses (N = 469,803).

| Characteristic | Number (%) |
| --- | --- |
| Men | 215,168 (45.8) |
| Women | 254,635 (54.2) |
| Mean Age (SD) | 56.5 (8.1) |
| Ancestry |  |
| White | 442,880 (94.3) |
| Mixed | 2,723 (0.6) |
| Asian | 9,099 (1.9) |
| Black | 7,277 (1.5) |
| Chinese | 1,455 (0.3) |
| Other | 4,173 (0.9) |
| Unknown | 2,196 (0.5) |

### Population-based odds ratios separate pathogenic and benign variants in many phenotypes

Among the 56,144 rare nonsynonymous variants we observed, we calculated population-based odds ratios for 5,737 variants (affecting 359,687 participants collectively) with sufficient biobank case data to calculate a valid corrected odds ratio (see **Methods**). The median odds ratio for a ClinVar pathogenic or likely pathogenic (P/LP) variant was 30.3 [IQR: 10.5 - 59.3], while the median odds ratio for a benign or likely benign (B/LB) variant was 1.1 [0.8 - 1.9], across all phenotypes. These figures vary across phenotypes, sometimes significantly. For cardiomyopathy-related phenotypes, odds ratios for pathogenic variants were particularly high, with phenotype-level median odds ratios for P/LP variants ranging from 125.1 (dilated cardiomyopathy) to 262.1 (hypertrophic cardiomyopathy) (**Figure 1A**). Some cancer related phenotypes had relatively lower odds ratios for damaging variants: breast/ovarian cancer had a median odds ratio for P/LP variants of 10.5, while Li-Fraumeni syndrome had a median of 8.3. For many phenotypes, B/LB variants had median odds ratios close to 1, with familial hypercholesterolemia and breast/ovarian cancer having a median odds ratio of exactly 1. Notably, for 11 phenotypes, the difference between the median odds ratios of P/LP and B/LB variants was statistically significant (one-sided Mann-Whitney U p < 0.05). The distribution of odds ratios for these phenotypes stratified by ClinVar status is shown in **Figure 1B**.

**Figure 1:**
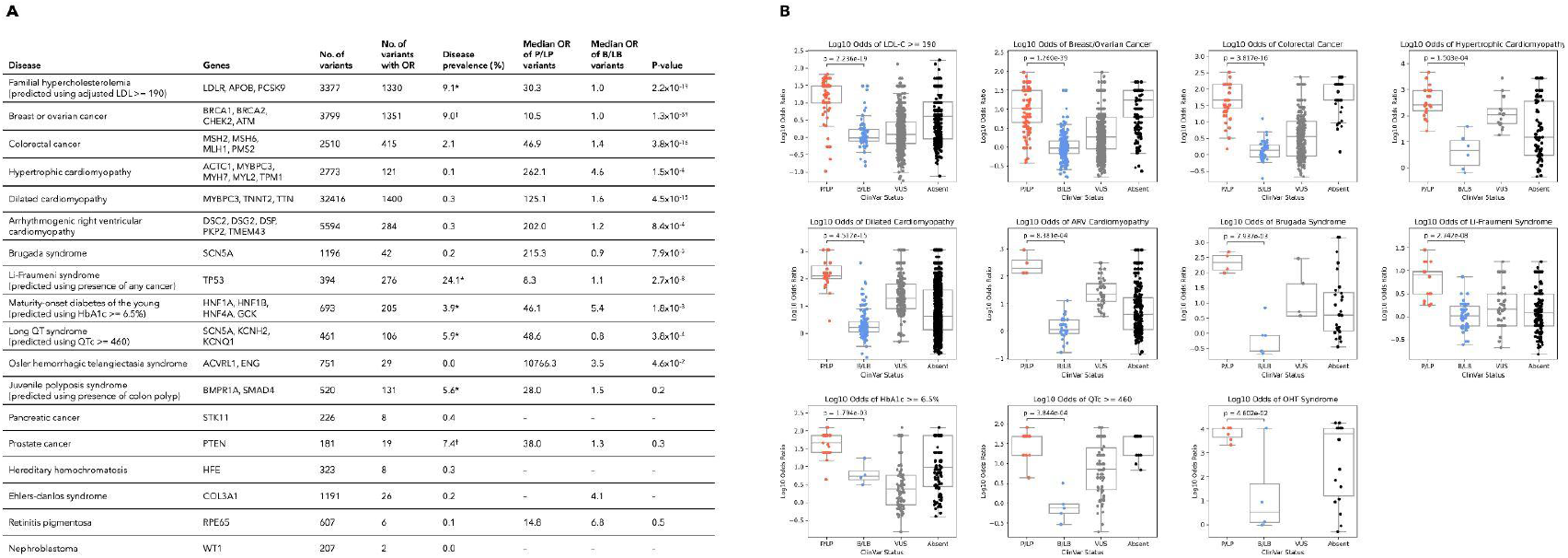
Population-based odds ratios separate pathogenic and benign variants across many phenotypes. **[A]** Phenotype-level information including genes considered, number of variants observed, number of variants observed for which an odds ratio could be calculated, cohort disease prevalence, median odds ratio of ClinVar pathogenic and benign variants, and one-sided Mann-Whitney U p-values between pathogenic and benign variant odds ratios. *Prevalence described uses the phenotypic encodings considered for participants to predict the presence of each disorder (*e*.*g*., prevalence of any cancer for Li-Fraumeni syndrome). ^†^Prevalence is based on participants of a single sex. **[B]** The distribution of log_10_ odds ratios by ClinVar status in 11 phenotypes with statistically significant separation of pathogenic and benign variants (one-sided Mann-Whitney U p < 0.05).

Next, we validated the discriminatory capacity of population-based odds ratios to classify pathogenic and benign variants. Across all phenotypes, we found that odds ratios were able to accurately classify existing ClinVar pathogenic and benign variants (AUC = 0.94). At the phenotypic level, AUCs ranged between 0.92 and 1.00 among 8 phenotypes where there were at least 25 ClinVar variants with either pathogenic (P/LP) or benign (B/LB) classifications with odds ratios (**Supplementary Figure 1**). We also calculate the optimal odds ratio thresholds at which specificity and sensitivity are maximized (calculated using Youdan’s Index, the most upper left point on ROC curves, which can also be interpreted as the point at which global LR+ is maximized), which can be used to dichotomously classify variants for each phenotype (**Supplementary Table 4**).

### Systematically calibrating evidence thresholds for population-based odds ratios

We sought to measure the strength of evidence provided by population-based odds ratios to align with the qualitative levels (‘supporting’, ‘moderate’, ‘strong’, ‘very strong’) described in the ACMG/AMP SVI guidelines, in order to accelerate the use of this information in variant interpretation. Using a local posterior-probability based approach described in **Methods**, we calibrate population-based odds ratios and define evidence thresholds for each phenotype with sufficient clinical data. Among 8 phenotypes with at least 25 pathogenic or benign ClinVar variants, the highest level of evidence reached was ‘very strong’ for 4 phenotypes (colorectal cancer, maturity-onset diabetes of the young, hypertrophic cardiomyopathy, arrhythmogenic right ventricular cardiomyopathy), ‘strong’ for 3 phenotypes (familial hypercholesterolemia, breast/ovarian cancer, Li-Fraumeni syndrome), and ‘moderate’ for 1 phenotype (dilated cardiomyopathy) (**Figure 2A**). Notably, we found that evidence thresholds for many phenotypes varied considerably from one another, demonstrating the utility of calibration at the phenotype level. This challenges the uniform application of specific odds ratio thresholds being treated as ‘strong’ evidence across all phenotypes, as described in the ACMG/AMP PS4 criterion. Phenotypes such as hypertrophic cardiomyopathy and arrhythmogenic right ventricular cardiomyopathy required higher odds ratios to reach the same level of evidence, while phenotypes such as familial hypercholesterolemia and breast/ovarian cancer required lower odds ratios to reach the same level of evidence. Evidence thresholds at the phenotypic level are presented in **Table 2**, and bootstrapped results and confidence intervals are presented in **Supplementary Figure 2A**.

**Table 2:**
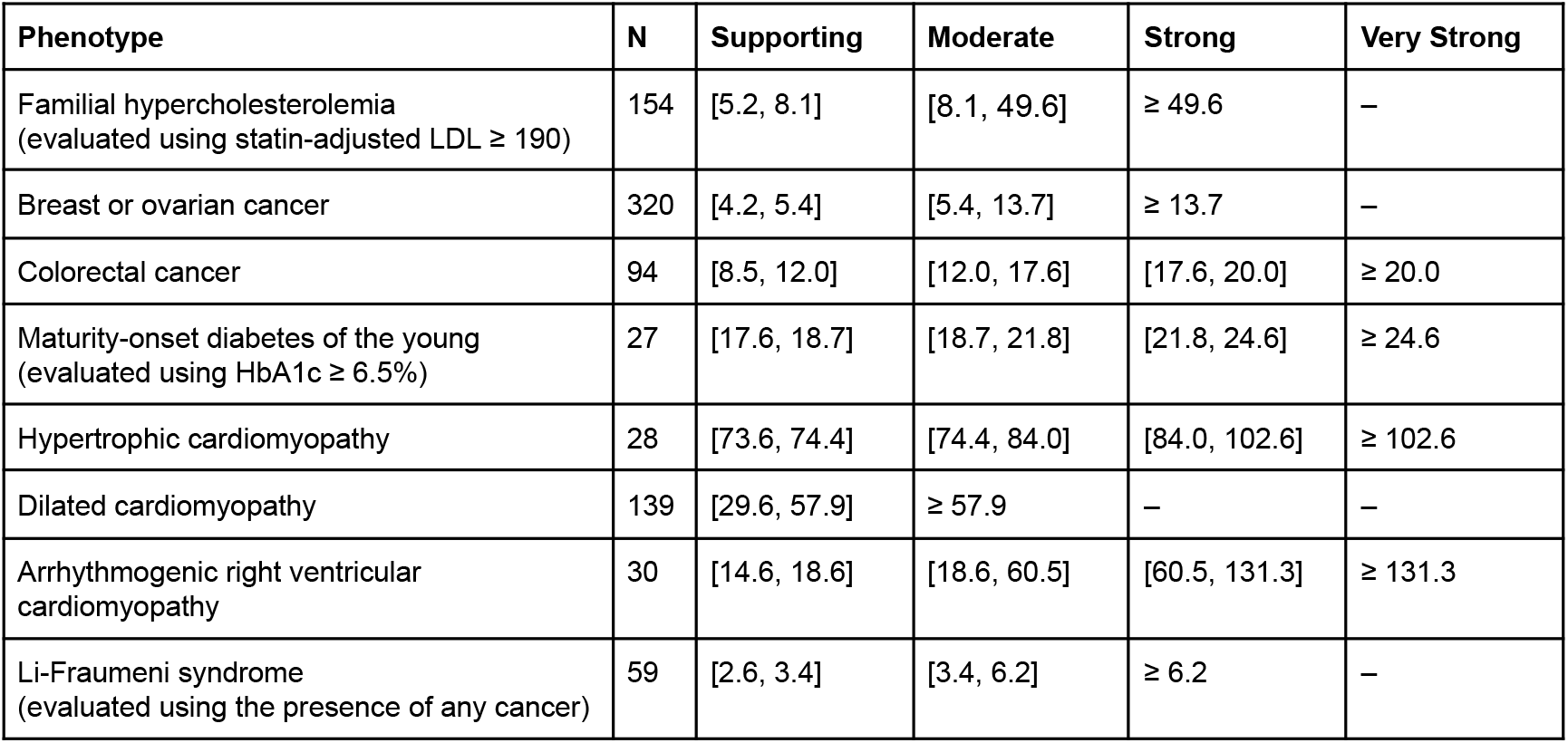
Estimated odds ratio evidence intervals at the phenotypic level. We calculated odds ratio intervals that correspond to different ACMG/AMP evidence strength levels for 8 phenotypes that had at least 25 pathogenic or benign ClinVar variants, so that there would be sufficient data for calibration. A dash indicates that the specified level of evidence was not reached. N represents the number of ClinVar pathogenic and benign variants considered for calibration in each phenotype.

**Figure 2:**
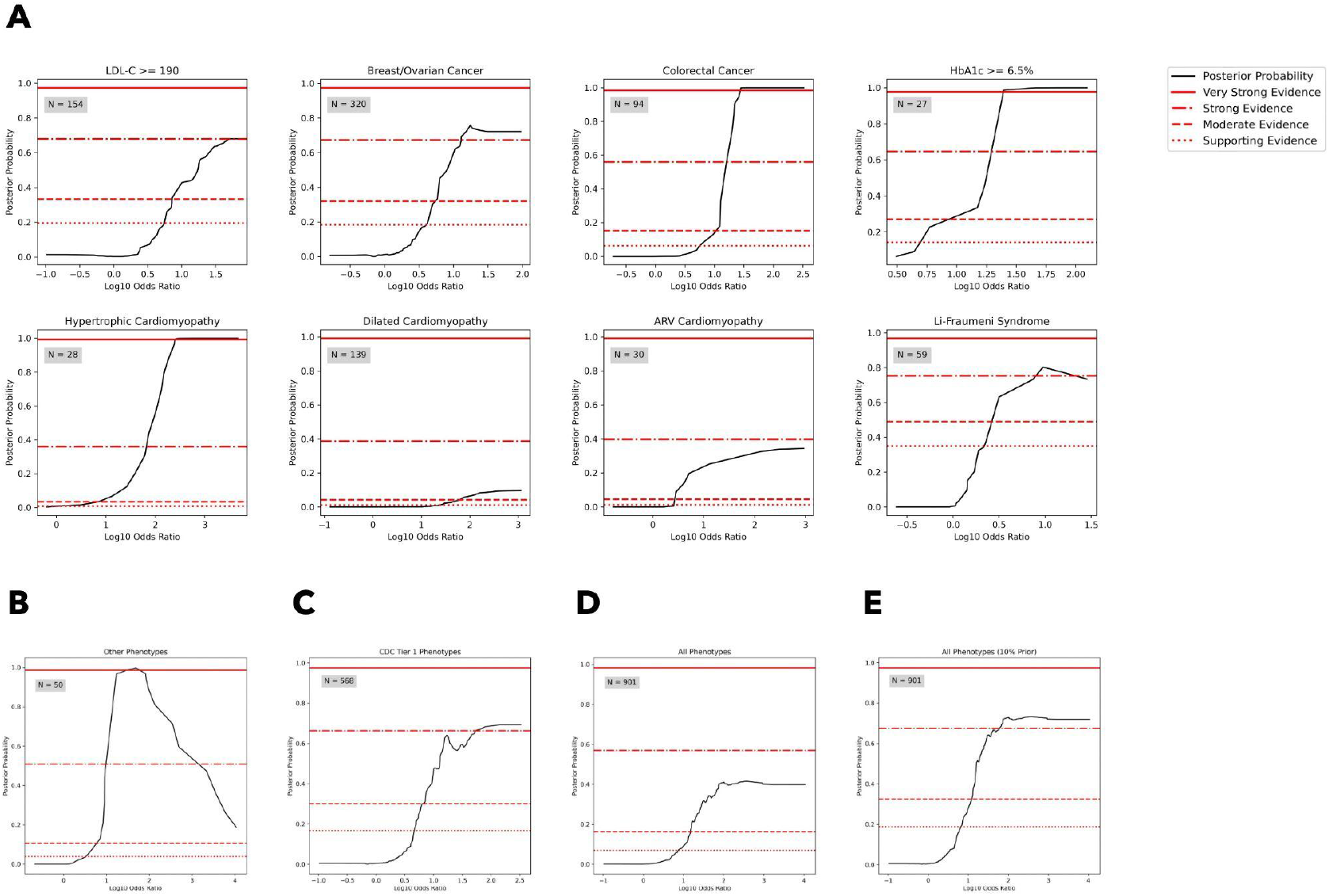
Posterior probability curves for select phenotypes and aggregations. **[A]** Posterior probability curves for variants in 8 phenotypes with at least 25 pathogenic or benign ClinVar variants available for evidence calibration. **[B]** Posterior probability curve for variants in phenotypes with fewer than 25 pathogenic or benign ClinVar variants, in aggregate. **[C]** Posterior probability curves for variants in phenotypes associated with CDC Tier 1 genes, in aggregate. **[D]** Posterior probability curves for variants in all phenotypes, in aggregate. **[E]** Posterior probability curves for variants in all phenotypes, in aggregate, using a prior probability of pathogenicity of 10%. Three SD Gaussian smoothing was applied to all plots uniformly. Threshold levels used (supporting, moderate, strong, and very strong, from bottom to top) vary by plot based on the estimation of distinct priors for each phenotype and aggregation, as described in **Methods**.

Beyond phenotype-level calibration, we also performed three aggregate calibrations across multiple phenotypes: (1) using phenotypes with fewer than 25 pathogenic or benign ClinVar variants, (2) using ‘CDC Tier One’ genes, and (3) using all phenotypes. For 10 phenotypes with fewer than 25 pathogenic or benign variants reached very strong evidence, phenotypes associated with CDC tier one genes (familial hypercholesterolemia, breast/ovarian cancer, colorectal cancer) reached strong evidence, and all phenotypes in aggregate reached moderate evidence (**Figure 2B-D**). These strength estimates are based on prior probabilities of disease that are developed using population sequencing data, and are consequently lower than disease priors previously used for calibration, leading to conservatively lower strengths of evidence (see **Methods**). When applying a commonly used prior of 10%, we find that all 18 phenotypes in aggregate reach strong evidence (**Figure 2E**). Evidence thresholds for these aggregations are presented in **Table 3**, and bootstrapped results and confidence intervals are presented in **Supplementary Figure 2B-E**.

**Table 3:** Estimated odds ratio evidence intervals for aggregations. We calculated odds ratio intervals that correspond to ACMG/AMP evidence strength levels for three aggregations across multiple phenotypes: phenotypes with fewer than 25 pathogenic or benign ClinVar variants, phenotypes associated with CDC Tier 1 conditions, and across all phenotypes. Strength levels for all phenotypes in aggregate were calculated using both a population-based prior (the default in our calculations) and a prior of 10%, a commonly used value. A dash indicates that the specified level of evidence was either not reached. N represents the number of ClinVar pathogenic and benign variants considered for calibration in each aggregation of phenotypes.

| Aggregation | N | Supporting | Moderate | Strong | Very Strong |
| --- | --- | --- | --- | --- | --- |
| Phenotypes with fewer than 25 pathogenic or benign ClinVar variants | 50 | [9.1, 9.6] | [9.6, 12.6] | [12.6, 16.1] | $\geq 16.1$ |
| Phenotypes associated with CDC Tier 1 conditions | 568 | [4.8, 6.9] | [6.9, 60.3] | $\geq 60.3$ | – |
| All phenotypes | 901 | [7.5, 14.5] | $\geq 14.5$ | – | – |
| All phenotypes (10% prior) | 901 | [6.7, 12.0] | [12.0, 40.9] | $\geq 40.9$ | – |

Using the full aggregate calibration thresholds based on a 10% prior across all 18 phenotypes we analyzed, among 53,926 VUS (including variants which are ‘uncertain’ or absent from ClinVar), we found that 456 VUS (0.8%) affecting 11,796 participants had supporting population evidence, 691 VUS (1.3%) affecting 9,596 participants had moderate population evidence, and 609 VUS (1.1%) affecting 2,754 participants had strong population evidence. Collectively, we found that 12,350 (2.6%) participants harbor a rare VUS with at least moderate evidence of pathogenicity in aggregate, highlighting the clinical value of population-based odds ratios. Using individual phenotype calibration thresholds instead, among 15,150 VUS in the 8 phenotypes we calibrated individually, we found that 172 VUS (1.1%) affecting 1,220 participants had supporting population evidence, 445 VUS (2.9%) affecting 2,085 participants had moderate population evidence, 266 (1.8%) VUS affecting 487 participants had strong population evidence, and 165 VUS (1.1%) affecting 337 participants had very strong population evidence of pathogenicity. We found that 2,909 (0.6%) participants harbor a rare VUS with at least moderate evidence of pathogenicity in these phenotypes.

When calculating odds ratios, ultra-rare variants have fewer observations and thus potentially less certainty around estimates for individual variants. To determine whether ultra-rare variants provide the same clinical certainty as a class when compared with variants with higher frequencies, we conducted a sensitivity analysis. Using a prior of 10%, we found that singletons – in this case, variants present in one participant who has an associated disorder – as a class provide ‘very strong’ evidence, as do variants present in 2 or 3 participants, though there is variance in these estimates at very high odds ratio values (**Supplementary Figure 3A**). More generally, we found that variants with at most 10 participants with the associated disorder reached the same level of evidence as variants with more than 10 participants (**Supplementary Figure 3B**). Furthermore, for singletons in particular, we found that 94.8% of singletons with an odds ratio of at least 5 with a decisive ClinVar classification were pathogenic (**Supplementary Figure 3C**). Notably, 93.9% of singletons with an odds ratio of at least 5 also had a lower 95th percentile confidence bound greater than 1, highlighting that even variants that only appear in one case can provide robust evidence.

### Comparing outcomes for participants with VUS with high odds ratios versus pathogenic variants

We sought to identify potential differences in clinical outcomes between participants with high odds ratio VUS (including variants which are ‘uncertain’ or absent from ClinVar) and participants with high odds ratio P/LP variants, using survival analysis. **Figure 3A** compares longitudinal clinical outcomes in *MSH2* and *MSH6*, two genes associated with colorectal cancer, at various odds ratio thresholds (5, 10, 15, and 30). Notably, despite being related parts of the same mismatch repair complex, VUS in *MSH2* must reach a much higher OR threshold in order to reach parity with similar P/LP variants in clinical outcomes, compared with VUS in *MSH6*, highlighting potential benefits of evaluating population evidence at the gene level. Beyond colorectal cancer, we observed large differences between genes related to the same phenotype, including *ATM* and *BRCA2* for breast cancer and *MYH7* and *MYBPC3* for hypertrophic cardiomyopathy, further motivating calibration at the gene level. We report calibration thresholds for 9 genes with at least 25 ClinVar pathogenic and benign variants in **Supplementary Table 5**.

**Figure 3:**
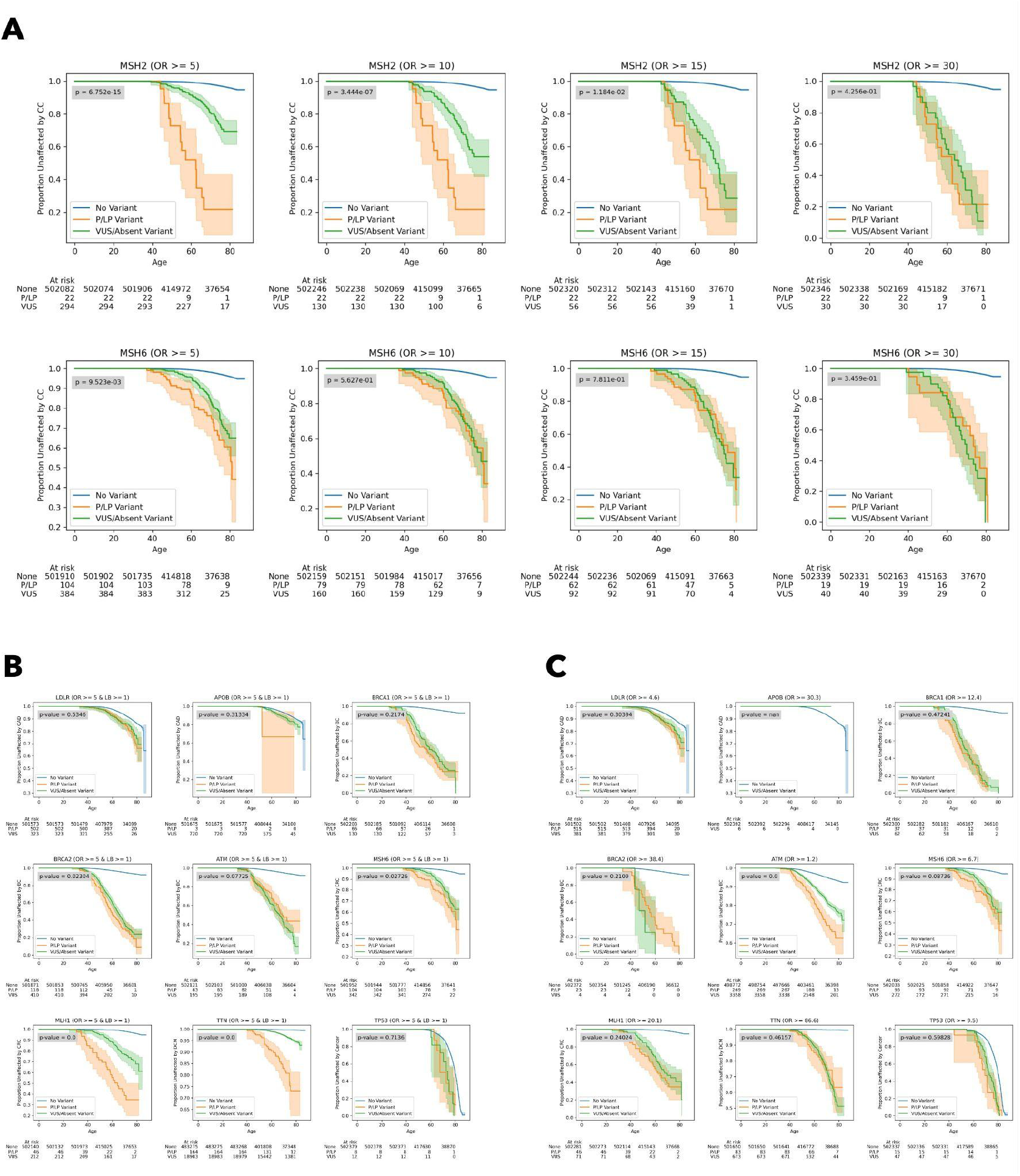
Comparing Kaplan-Meier curves for participants with high odds ratio VUS versus participants with high odds ratio pathogenic variants. Survival curves were generated using a Kaplan-Meier estimator, and shaded regions represent 95% confidence intervals.”At risk” counts describe the number of participants considered in each of the three groups at different ages. Logrank p-values between P/LP and VUS survival curves are noted in each plot. **[A]** Survival analysis of participants with no variants, P/LP variants, and VUS/absent variants in MSH2 and MSH6. Each column considers variants above an odds ratio threshold ranging from 5 to 30. **[B]** Survival analysis of participants with no variants, P/LP variants with odds ratios ≥ 5/lower bounds ≥ 1, and VUS/absent variants with odds ratios ≥ 5/lower bounds ≥ 1 in each of 9 genes which had at least 25 ClinVar pathogenic and benign variants. **[C]** Survival analysis of participants with no variants, P/LP variants with odds ratios ≥ optimal threshold, and VUS/absent variants with odds ratios ≥ ‘optimal threshold’ in each of 9 genes which had at least 25 ClinVar pathogenic and benign variants. Note that optimal thresholds refer to the thresholds like those described in **Supplementary Table 3** but at the gene level.

When applying the ACMG/AMP recommended threshold values for the application of PS4 population evidence (variants with an odds ratio ≥ 5 and a lower 95% confidence bound ≥ 1), we found that outcomes among participants with VUS were not significantly different from those with P/LP variants in some genes. Two genes associated with colorectal cancer: *MSH6* (logrank p = 0.03) and *MLH1* (logrank p = 1.4×10^−8^), as well as *BRCA2* (logrank p = 0.02) and *TTN* (logrank p = 2.1×10^−31^) still had differences between clinical outcomes for patients with VUS versus P/LP variants (**Figure 3B**). This indicates that PS4 evidence developed using this approach based on current guidelines – in aggregate – may be as strong as P/LP annotations in some, but not all, of the genes we evaluated.

We next analyzed outcomes in the same genes using optimal odds ratio thresholds (values at which specificity and sensitivity are maximized) at the gene level (**Supplementary Table 4**), and find that there is no significant difference in outcomes between participants that have VUS and P/LP variants with an odds ratio greater than the optimal threshold in all genes except *ATM* (logrank p = 6.4×10^−7^) and *APOB* (no participants have P/LP variants with OR ≥ optimal threshold) (**Figure 3C**). When compared with the ACMG/AMP PS4 recommended odds ratio thresholds for PS4 evidence, optimal threshold values are sometimes higher, and result in similar clinical outcomes between patients with P/LP and VUS variants in more genes (7 of 9 versus 5 of 9).

### Correlation between population, computational, and functional evidence

We analyzed the correlation between odds ratios (PS4/population evidence), REVEL scores (PP3/BP4/computational evidence), and functional scores (PS3/BS3/functional evidence) in 3 genes where these data sources were all available (*LDLR, BRCA1, MSH2*). Interestingly, we found that odds ratios are moderately correlated with REVEL scores in *LDLR*, but not in *BRCA1* and *MSH2* (**Figure 4A**), and that odds ratios are moderately correlated with functional scores in *LDLR* and *BRCA1*, but not in *MSH2* (**Figure 4B**). We note that *MSH2* requires a much higher odds ratio than *LDLR* and *BRCA1* to reach the same level of evidence, which may contribute to the lack of correlation with other forms of evidence, given that the majority of variants we evaluated in *MSH2* were below that threshold.

**Figure 4:**
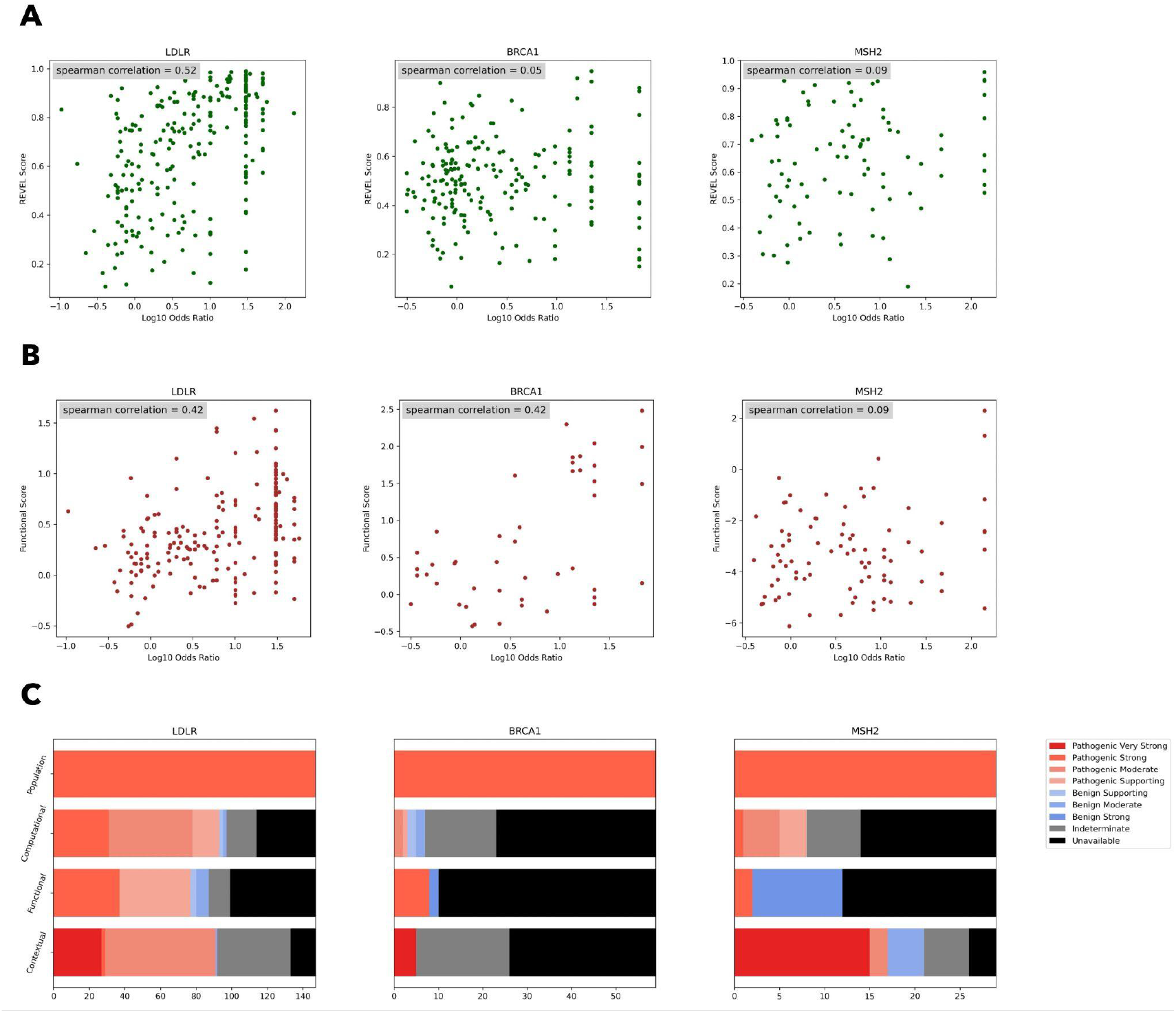
Correlation between population evidence and computational or functional evidence. **[A]** Spearman correlation between log_10_ odds ratios and REVEL computational predictions of variant pathogenicity. **[B]** Spearman correlation between log_10_ odds ratios and functional estimates of variant impact from experimental assays. **[C]** Number of variants with different forms of evidence (computational, functional, contextual) among variants that already have strong population evidence of pathogenicity.

We then sought to identify how many VUS in these genes might have sufficient evidence to be ‘pre-classified’ as P/LP, making use of population, computational, and functional evidence, as well as contextual evidence from previously classified pathogenic variants, which has been shown to be potentially underused.^12^ We evaluate each of these forms of evidence for variants that meet the threshold for strong population evidence of pathogenicity (≥ 6.6 for *LDLR*, ≥ 23.2 for *BRCA1*, and ≥ 24.0 for *MSH2*), as described in the **Methods**. The number of variants in *LDLR, BRCA1*, and *MSH2* with each form of evidence is shown visually in **Figure 4C**.

We combine these forms of evidence using the Bayesian framework based point system, noting that a comprehensive evaluation of all available forms of evidence would be required for a diagnostic variant interpretation. Using this approach, we are able to ‘pre-classify’ 60 VUS across *LDLR, BRCA1*, and *MSH2* (80% in *LDLR*), affecting 245 participants, as P/LP (points ≥ 6) (**Supplementary Figure 4**). Notably, in *LDLR*, 72.2% of VUS with strong population evidence of pathogenicity also have sufficient complementary evidence to be potentially classified as pathogenic. These VUS with sufficient evidence for classification represent 12.4% of all rare VUS in *LDLR* observed in participants, suggesting that there may be a high potential yield of variants that could be reclassified in some genes when well-calibrated functional and population evidence are available.

## Discussion

Collectively, many individuals harbor a rare VUS in an actionable disease gene, and this framework can provide information to significantly accelerate the interpretation of these challenging variants. Our results demonstrate that case enrichments within large population cohorts can broadly provide meaningful clinical evidence for rare coding VUS. This approach can streamline the generation of new information for variant assessment across a range of clinically actionable phenotypes, including rare and common disorders. We found that population-based odds ratios collectively reached ‘strong’ evidence when applying the Bayesian adaptation of the ACMG/AMP framework when applying a commonly used prior of 10%.^13^ At the phenotypic level, population odds ratios reached at least ‘moderate’ of evidence in all 8 phenotypes with sufficient numbers of variants for calibration, with multiple phenotypes reaching ‘strong’ and ‘very strong’ evidence.

### Calibration of evidence at the phenotype and gene level

We calibrated population evidence at the phenotype and gene level, defining odds ratio thresholds which are considered ‘supporting’, ‘moderate’, ‘strong’, or ‘very strong’ evidence of pathogenicity. Our approach involved calibrating and calculating prior probabilities of pathogenicity for each phenotype and gene from a biobank dataset, in contrast to previous calibration methods that use a single threshold and prior probability across all genes.^13,14^ The considerations and tradeoffs in different approaches to calculating prior probabilities are discussed extensively in **Methods**. We used these values to calculate local likelihood ratios for each gene and phenotype. Given the wide variability we observed in evidence thresholds and the prevalence of pathogenic variants among different genes and phenotypes, this approach may be valuable in future calibration efforts when sufficient data is available more broadly. It is also likely to be useful for informing the specialized work of VCEPs to determine how to apply various evidence strength thresholds for specific genes and sub-types of disorders.

We highlighted an example in the mismatch repair complex, where we observed that variants in *MSH2* and *MSH6* had different odds ratio thresholds to reach the same clinical certainty. These differences at the gene level may be statistical artifacts related to different numbers of variants with odds ratios, numbers of pathogenic and benign variants, or differences in accuracy of prior clinical classifications. This may also have a biological basis; while Msh2 and Msh6 physically interact and form the Muta complex, each subunit contributes uniquely to the repair process. Msh2 plays a central role in stabilizing the complex, while Msh6 provides the specificity for recognizing particular types of DNA mismatches. There are also existing known differences in effect sizes of protein truncating variants, for example, such variants in *BRCA1* or *BRCA2* are much more likely to lead to cancer when compared with protein truncating variants in *ATM*.^15^

Calibrating the strength of evidence at the gene and phenotype level also has limitations, as there are currently low numbers of ClinVar pathogenic and benign variants in many genes, which can naturally increase variance around calibration thresholds. Case ascertainment and available case data may differ substantially across genes, leading to varying levels of clinical certainty for pathogenic and benign variants in different genes. This is also confounded by biological factors, such as varying phenotypic effect sizes, incomplete penetrance, or stochasticity related to phenotypic effects (*e*.*g*., for endophenotypes like LDL-C leading to myocardial infarction or DNA damage repair competency leading to cancer). There may also be specialized biological modeling about variant types (*e*.*g*., cysteine residues within *LDLR* class A repeats) which could inform these evidence models.

### Approaches to automating components of variant interpretation

Methods to automatically generate structured sources of diagnostic evidence can expedite variant assessment and prioritize the most promising variants for re-assessment. Notably, when combining four forms of evidence which we automatically generated (population, computational, functional, contextual), we found that 72.2% of VUS in *LDLR* with strong or very strong population evidence also have sufficient complementary evidence to be potentially classified as pathogenic, and that these VUS with sufficient evidence represent 12.4% of all rare VUS in *LDLR* seen in participants. Broadly, among participants with a rare variant in the genes we analyzed that reached strong or very strong evidence, 4.3% (N = 2,456) have a VUS or variant absent from ClinVar with strong population evidence that could be informative about increased risk of a related disorder. As functional assay data for additional genes are developed, and computational scores evolve, combining these sources of evidence can help dramatically scale variant interpretation. We note that broad application of this framework will require a careful review of VCEP classification criteria (*e*.*g*., evidence type PM5 should not be applied when classifying variants in *BRCA1* or *BRCA2*).

### Limitations and future directions

In populations other than those most highly represented in the UK Biobank, there are an insufficient number of participants with rare missense variants to make robust estimates of risk. Therefore, we note that our estimates may not be generalizable to those other populations or those not ascertained during adulthood, and future work may estimate population-specific variant effects. Additionally, while our analysis focuses on a subset of actionable genes that are some of the most commonly screened in diagnostic settings, future work may analyze a broader set of genes. Given small variant counts at the gene level, we caution that gene-level calibration may be data-constrained, and that statistical estimates will become more powerful as biobank sizes grow. We provide higher confidence calibration thresholds in our fully aggregated calibration across all 11 genes we analyzed (**Supplementary Table 4**).

To remain consistent with ACMG/AMP guidelines and commonly used standards developed by variant curation expert panels (VCEPs), we use an odds ratio to represent enrichment of disease at the population level for individual variants. Future work may evaluate whether other representations of population evidence (*e*.*g*., using other statistical measures or models) can achieve better performance. We note that odds ratio estimates can be confounded by low variant allele counts or population structure. Separately, because we have focused on rare variants in genes where coding variants are known to have a substantial effect, we presume that these variants are likely to be truly causal. In rare cases, it is possible for a variant we analyzed to be tagged by a more common coding variant with functional effect, though this is unlikely to impact our calibration efforts which were aggregated at the gene level. Additionally, many complex disorders are attributable to other complex genetic, behavioral, or environmental factors, in addition to their established monogenic contributions.

### Conclusion

In summary, this analysis presents a comprehensive approach to assess the impact of germline variants using endophenotypic and disease risk data from a national biobank. For the set of genes we analyzed, we calculated variant-level odds ratios and calibrated strengths of evidence, then used these to identify VUS which can potentially be re-classified as pathogenic. By highlighting the utility of biobank data and calibrating it, we hope that this form of population evidence can be adopted to inform variant interpretation broadly.

## STAR ★ Methods

### Key resources table

| REAGENT or RESOURCE | SOURCE | IDENTIFIER |
| --- | --- | --- |
| **Deposited data** |  |  |
| UK Biobank WES dataset | Backman et al.<sup>16</sup> | https://www.ukbiobank.ac.uk |
| UK Biobank phenotype dataset | Bycroft et al.<sup>17</sup> | https://www.ukbiobank.ac.uk |
| LDLR base editing dataset | Ryu et al.<sup>18</sup> | Supplementary Tables |
| BRCA1 SGE dataset | Findlay et al.<sup>19</sup> | Supplementary file 2 |
| MSH2 DMS dataset | Jia et al.<sup>20</sup> | mavedb:00000050-a-1 |
| ClinVar | NCBI | v04/06/2023 |
| **Software and algorithms** |  |  |
| bcftools | SAMtools | v1.15.1 |
| Swiss Army Knife | DNAnexus Apps | v4.9.1 |
| Variant Effect Predictor | Ensembl (McLaren et al.<sup>21</sup>) | v108 |
| Python | Python Software Foundation | v3.8.5 |
| SciPy | Virtanen et al.<sup>22</sup> | v1.7.3 |
| lifelines | Davidson-Pilon et al.<sup>23</sup> | v0.23.9 |
| Pandas | McKinney et al.<sup>24</sup> | v1.1.4 |
| NumPy | Harris et al.<sup>25</sup> | v1.21.1 |
| Matplotlib | Hunter et al.<sup>26</sup> | v3.3.2 |
| Seaborn | Waskom et al.<sup>27</sup> | v0.11.0 |

## Resource availability

### Lead contact

Requests should be directed to and will be fulfilled by the lead contact, Christopher A. Cassa

### Materials availability

This study did not generate new unique reagents.

### Experimental model and subject details

No experimental models or novel subjects were utilized or collected as part of this publication.

## Method details

### Study participants

The UK Biobank is a prospective cohort of over 500,000 individuals recruited between 2006 and 2010, ages 40-69.^28^ Among 469,803 participants with whole exome sequencing data, we evaluated rare non-synonymous variants present in genes related to 18 actionable disorders, as shown in **Figure 1A**. We excluded participants who either had missing phenotypic data, who had more than one rare (AF <= 0.1%) non-synonymous variant across the set of genes related to each phenotype, or who had a rare non-synonymous homozygous variant.

### *Variant inclusion, quality control, and annotations* from exome sequencing data

#### Sequencing and transcripts

Whole exome sequencing (WES) was performed for UK Biobank participants, as previously detailed.^10^ Analysis was conducted on the UK Biobank DNAnexus Research Analysis Platform (https://ukbiobank.dnanexus.com). We extracted gene-level VCF files from WES pVCFs and normalized to flatten multi-allelic sites using bcftools (v1.15.1) and the Swiss Army Knife application (v4.9.1).^29^ Exon coordinates were identified from MANE transcripts, with an additional 5 nucleotides retained upstream and downstream of each coding region to capture splice-site variants.

#### Quality control

Variants in low complexity regions, segmental duplications, or other regions known to be challenging for next generation sequencing alignment or calling were removed from analysis (NIST GITB difficult regions),^30^ as were variants with an alternate allele frequency greater than 0.1% in the UK Biobank cohort.^10^ Further filtering removed variants in which more than 10% of samples were missing genotype calls. To mitigate differences in sequencing coverage between individuals who were sampled at different phases of the UK Biobank project, variants were only retained in the final set if at least 90% of their called genotypes had a read depth of at least 10.

#### Annotations

The canonical functional consequence of each variant was calculated using Variant Effect Predictor (v108).^21^ Non-coding variants outside of essential splice sites were not considered in the analysis, and non-synonymous coding variants were included with any of the following canonical consequences:”splice_acceptor_variant”,”splice_donor_variant”,”stop_gained”,”frameshift_variant”,”stop_lost”,”start_lost”,”missense_variant”,”inframe_insertion”,”inframe_deletion”. REVEL scores were aggregated from all available transcripts and annotated if available in at least one transcript.

### Clinical endpoints and endophenotypes

Primary clinical endpoints were specific to each condition. For familial hypercholesterolemia, maturity-onset diabetes of the young, and long QT syndrome, endophenotype values above a threshold were used as a proxy for disease cases. Adjusted LDL-C ≥ 190 mg/dL, HbA1c ≥ 6.5%, and QTc ≥ 460 were used for familial hypercholesterolemia, maturity-onset diabetes of the young, and long QT syndrome, respectively. Estimated untreated (adjusted) LDL-C levels were derived using adjustments for lipid-lowering therapies, as applied previously.^31^ Additionally, presence or absence of any cancer was used as a proxy for Li-Fraumeni syndrome, and presence of a colon polyp was used as a proxy for juvenile polyposis syndrome. Note that these ‘cases’ derived from alternative data may not be considered cases in the traditional sense; however, their use allows for predicting variant impact even in the absence of direct data on disorders. For all other disorders, cases were ascertained using presence or absence of the specific disorder. Case definitions were developed in the UK Biobank using a combination of self-reported data confirmed by trained healthcare professionals, hospitalization records, and national procedural, cancer, and death registries, when applicable. Age at event was estimated based on the listed date of event, when available, and derived using participant birth dates when not directly provided. Cases without any form of age at event information were excluded.

### Calculating variant-level odds ratios from population cohort data

Variant-level odds ratios were calculated using the aforementioned endophenotype and disorder case data, where for a non-synonymous variant *v* the”population-based odds ratio”

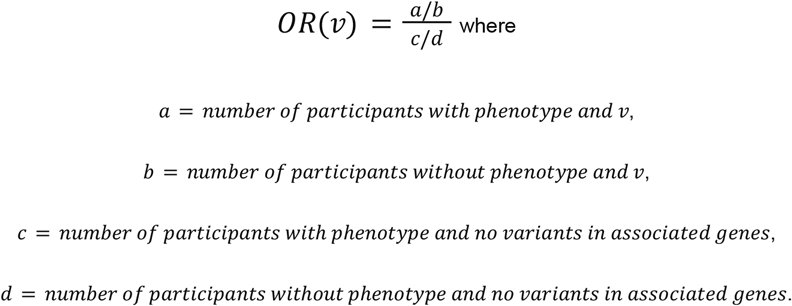

Haldane-Anscombe correction (adding 0.5 to all cells *a, b, c, d* in the contingency table) was applied to allow for the calculation of odds ratios when no false positives existed, while also reducing the bias of the estimator. Variants for which *a* was 0 and the corrected odds ratio was ≥ 1 were not included in analyses.

### ClinVar clinical assertions from diagnostic laboratories

ClinVar summary assessments were extracted from the tab delimited ClinVar variant summary file released on 04/06/2023.^32^ We grouped ‘pathogenic’, ‘likely pathogenic’, and ‘pathogenic/likely pathogenic’ classifications as ‘P/LP’ collectively, and grouped ‘benign’, ‘likely benign’, and ‘benign/likely benign’ classifications as B/LB. Importantly, we used the term VUS inclusively to capture all variants with an inconclusive classification: including”uncertain significance,”“conflicting interpretations of pathogenicity,” and”not provided”.

### Calibrating population evidence of pathogenicity (PS4)

#### Estimating the prevalence of pathogenic variants

We considered two methods for estimating the prevalence of pathogenic variants, or the prior probability of pathogenicity: 1) using population data from the UK Biobank and 2) using clinical data from ClinVar. In the first approach, we estimate the prior probability of pathogenicity in a gene *g* as

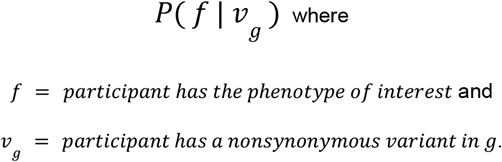

Then, priors at the phenotypic level can be calculated as the weighted average

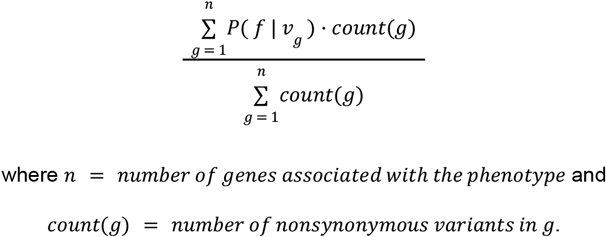

A similar weighted average can be used to aggregate phenotype-level priors into an overall prior for all phenotypes considered, or some subset of phenotypes. Often, in previous studies, a single overall prior has been calculated for calibration across all genes in a dataset; however, we contend that there is a benefit to stratifying at the phenotype and gene level given how widely priors can differ at these levels.^13,14^ We report prior probabilities of pathogenicity generated using population data in **Supplementary Table 1, Supplementary Table 2**, and **Supplementary Table 3**. These priors generally closely track disease prevalence, with deviations in some phenotypes. Notably, the aggregated prior across phenotypes associated with CDC tier one genes was 8.6%, similar to the 10% prior assumed in *Tavtigian et al*.^13^ The aggregate prior across all phenotypes was lower, at 2.8%.

In the second approach, we estimate the prior probability of pathogenicity in a gene *g* as

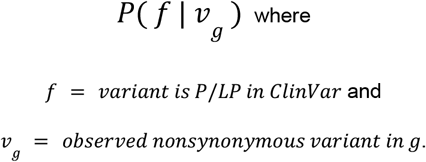

Weighted averages can be calculated for phenotype-level and further aggregated prevalence values of pathogenic variants, as described earlier. We report priors generated using clinical data in **Supplementary Table 1, Supplementary Table 2**, and **Supplementary Table 3**, next to the priors generated using population data. Compared to priors based on population data, these priors were generally higher in low disease prevalence phenotypes and lower in high disease prevalence phenotypes. Notably, the aggregated prior across phenotypes associated with CDC tier one genes was 5.4%, similar to the 4.41% calculated in *Pejaver et al*.^14^ The aggregate prior across all phenotypes was lower, at 1.9%.

We caution that both of our methods to estimate pathogenic variant prevalence may underestimate true values. When using population case data from the UK Biobank, there are participants that are yet to develop a phenotype, and when considering the proportion of ClinVar P/LP variants among nonsynonymous variants in the UK Biobank, there are a large number of nonsynonymous variants observed in the UK Biobank that are not represented in ClinVar. For subsequent analysis, we use population-based priors, as some phenotypes we evaluated have very few ClinVar variants, and participants in the UK Biobank have been enrolled over a long period, so we are less likely to underestimate relevant values for developing these priors.

#### Statistical framework

The ACMG/AMP variant interpretation guidelines describe multiple levels of strength in favor of variant pathogenicity: supporting, moderate, strong, and very strong.^6^ These strength levels have been mapped to positive likelihood ratios (LR+) to quantify variant impact.^13^ To calibrate the strength of population evidence (which we model as continuous odds ratios) at the gene level, we use a sliding window method similar to that introduced in *Pejaver et al*. for the calibration of computational scores.^14^ For every population odds ratio *p*, we calculate a local likelihood ratio (lr+) using pathogenic and benign variants with odds ratios in the interval [*p* − *z, p* + *z*] for the lowest value of z such that {*v*| *OR*(*v*) ∈ [*p* − *z, p* + *z*]} contains at least 20% of all pathogenic and benign variants in a given gene excluding those for which odds ratios could not be calculated. The local likelihood ratio calculation is then

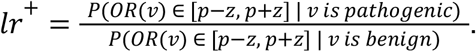

Given this and the prior probability of pathogenicity *a*, we can calculate the posterior probability of pathogenicity *b* for a variant by rearranging the following equation:

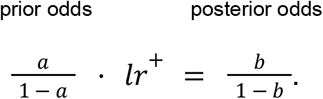

Next, to map supporting, moderate, strong, and very strong evidence thresholds to posterior probability thresholds for each gene and phenotype, we calculate suitable values for the odds of pathogenicity for very strong evidence (*O*_*PVst*_). These values were determined using the supplementary table from *Tavtigian et al*. so that the ACMG/AMP combining rules are generally satisfied and posterior probabilities reach the values of 0.9 and 0.99 for likely pathogenic and pathogenic classifications, respectively. O_PVst_ values are reported in **Supplementary Table 1, Supplementary Table 2**, and **Supplementary Table 3**, next to their respective prior probabilities. We then used the *O* _*PVst*_ values for each gene, phenotype, and further aggregation as the *lr* ^+^ variable in the equation above to calculate the very strong posterior probability threshold. For strong, moderate, and supporting evidence of pathogenicity, this process was repeated but using 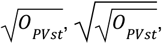 and 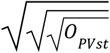, respectively, based on the assumption that evidence levels scale by powers of 2.

Finally, to identify intervals of population-based odds ratios that correspond to varying levels of evidence, we identify the minimum odds ratio at which the posterior probability threshold for an evidence level is crossed, then use linear approximation to estimate the odds ratio at which the exact threshold would be reached.

### Alternative approaches considered for calibration

Our calibration strategy follows a localized estimation approach as described previously. We considered this localized approach and a global approach which involves calculating likelihood ratios using all available variants. The primary disadvantage of a global approach is that only a single global threshold is considered, and any odds ratio value greater than that threshold would be considered to have the same strength of evidence, though this may not always be the case.^14^ Due to this challenge, we used a localized estimation approach, though we note that this also has limitations. Specifically, a local approach requires a large amount of data and may suffer from imprecision when the interval around a score used to calculate *lr*^+^ has to be very wide in order to include a sufficient number of variants for estimation. While the approach in *Pejaver et al*. maintained intervals wide enough such that 100 pathogenic or benign variants were always included, this approach was not always possible with population data at the gene level given the low numbers of pathogenic or benign variants in some phenotypes and genes. Instead, we used intervals wide enough such that 20% of pathogenic or benign variants are included, as we found this to work particularly well, although alternative approaches may use a different proportion or a constant number.

Estimating the prevalence of pathogenic variants, or the prior probability, can be substantially different depending on the context. For example, *Tavtigian et al*. assumed a prior of 10% given the context of identifying a pathogenic variant among a set of candidate variants from clinical genetic testing, and *Pejaver et al*. estimated a lower number, 4.41%, measured empirically using gnomAD as a reference set. For our analyses, as described previously, we used priors using population data from the UK Biobank due to their concordance with our approach, though we also calculated priors based on clinical data (using non-pathogenic UK Biobank variants as controls). We note that other approaches based on clinical diagnostic data may make use of other control datasets such as gnomAD for this purpose, and that there are alternative ways to calculate prior probabilities outside of these two methods (*e*.*g*., using functional assay data).

### Survival analysis

Survival analyses including Kaplan-Meier curves and logrank tests were performed using the Python lifelines survival analysis package (v0.23.9).^23^

### Conversion of evidence to the point system

To identify how many VUS and variants absent from ClinVar with population evidence might have sufficient evidence to be classified as P/LP, we considered multiple evidence sources and converted them to the semi-quantitative point system adaptation of the ACMG/AMP sequence variant interpretation framework.^33^ We interpreted variants with ORs greater than the strong threshold calibrated for the gene as having PS4 evidence (+4 points on the point scale). REVEL scores were converted to evidence bands (PP3/BP4) using thresholds calculated in *Pejaver et al*.,^14^ and functional scores were translated to PS3 (+4 points) or BS3 (−4 points) based on author recommended thresholds, with the exception of *LDLR*, where a threshold wasn’t provided and which we estimated by analyzing global LR+ with ClinVar as a truth set.^18–20^ Finally, contextual evidence from previously classified pathogenic variants was applied in the following manner: PVS1 (+8 points) for LOF variants annotated as high-confidence by LOFTEE,^34^ PS1 (+4 points) when a variant has a colocated P/LP variant encoding the same substitution and −4 points when a variant has a colocated B/LB variant encoding the same substitution (note that the benign equivalent is not an established ACMG/AMP criterion), and PM5 (+2 points) when a variant has a colocated P/LP variant encoding a different substitution and −2 points when a variant has a colocated B/LB variant encoding a different substitution (note that the benign equivalent is not an established ACMG/AMP criterion; PM5 was not applied to *BRCA1* in accordance with guidance from the VCEP).^35^

## Data Availability

All data produced are available online at:

https://doi.org/10.6084/m9.figshare.28776104.v1

## Acknowledgments

We are grateful to the UK Biobank and its participants who provided biological samples and data for this study, performed under UK Biobank application 41250 and Mass General Brigham IRB protocol 2020P002093. We gratefully acknowledge funding from NIH R01HG010372 (V.B., T.Y., L.B., C.C.), R01HG013350 (V.P.) and R56HG012681 (T.Y., C.C.), and the American Heart Association (24TPA1300072: T.Y., C.C.).

## Data availability

Variant-level odds ratios and the underlying case data used in their calculation are available at https://doi.org/10.6084/m9.figshare.28776104.v1 for all 18 phenotypes studied.

---

**Supplementary Table 1:** Prior probabilities of pathogenicity at the gene level. Prior probability of pathogenicity estimated based on population data (column 2) and clinical data (column 4) for each gene. The odds path for very strong evidence, O_PVst_, associated with each prior, is reported in adjacent columns.

| **Gene** | **Prior**<br>**(Population)** | **O<sub>PVst</sub>**<br>**(Population)** | **Prior**<br>**(Clinical)** | **O<sub>PVst</sub>**<br>**(Clinical)** |
| --- | --- | --- | --- | --- |
| LDLR | 19.3% | 127 | 20.1% | 120 |
| APOB | 9.2% | 398 | 0.7% | 12704 |
| PCSK9 | 4.5% | 1111 | 1.6% | 4660 |
| BRCA1 | 9.2% | 394 | 5.3% | 874 |
| BRCA2 | 9.7% | 365 | 5.9% | 749 |
| CHEK2 | 12.4% | 255 | 10.2% | 341 |
| ATM | 9.6% | 375 | 8.2% | 470 |
| MSH2 | 2.5% | 2450 | 3.1% | 1816 |
| MSH6 | 2.3% | 2859 | 6.7% | 629 |
| MLH1 | 3.0% | 1909 | 6.3% | 681 |
| PMS2 | 1.9% | 3605 | 6.0% | 743 |
| ACTC1 | 0.2% | 64521 | 0.0% | inf |
| MYBPC3 (Hypertrophic cardiomyopathy) | 0.3% | 36218 | 2.6% | 2343 |
| MYH7 | 0.2% | 75927 | 3.6% | 1500 |
| MYL2 | 0.3% | 47294 | 0.5% | 23814 |
| TPM1 | 0.1% | 181035 | 1.1% | 7524 |
| MYBPC3 (Dilated cardiomyopathy) | 0.2% | 57506 | 2.6% | 2343 |
| TNNT2 | 0.3% | 35647 | 2.0% | 3464 |
| TTN | 0.3% | 41090 | 0.6% | 17354 |
| DSC2 | 0.3% | 37517 | 0.4% | 32993 |
| DSG2 | 0.4% | 32605 | 0.6% | 18093 |
| DSP | 0.3% | 37095 | 1.6% | 4635 |
| PKP2 | 0.4% | 25541 | 2.2% | 2895 |
| TMEM43 | 0.3% | 37126 | 0.2% | 83340 |
| SCN5A (Brugada syndrome) | 0.2% | 74660 | 2.0% | 3342 |
| TP53 | 23.2% | 103 | 5.6% | 813 |
| HNF1A | 6.5% | 661 | 8.7% | 429 |
| HNF1B | 7.2% | 572 | 2.9% | 1982 |
| HNF4A | 6.3% | 685 | 3.4% | 1602 |
| GCK | 9.7% | 366 | 25.0% | 96 |
| SCN5A (Long QT syndrome) | 5.7% | 782 | 2.7% | 2205 |
| KCNH2 | 16.5% | 164 | 4.9% | 983 |
| KCNQ1 | 15.6% | 178 | 16.5% | 164 |
| ACVRL1 | 0.1% | 195834 | 4.6% | 1074 |
| ENG | 0.1% | 206486 | 1.6% | 4410 |
| BMPR1A | 5.0% | 961 | 0.4% | 29170 |
| SMAD4 | 5.8% | 764 | 0.4% | 32670 |
| STK11 | 0.3% | 48441 | 0.0% | inf |
| PTEN | 3.0% | 1909 | 8.3% | 462 |
| HFE | 0.2% | 81510 | 2.5% | 2508 |
| COL3A1 | 0.2% | 61579 | 1.4% | 5305 |
| RPE65 | 0.1% | 237321 | 8.6% | 440 |
| WT1 | 0.0% | 2352837 | 1.0% | 8981 |

**Supplementary Table 2:** Prior probabilities of pathogenicity at the phenotypic level. Prior probability of pathogenicity estimated based on population data (column 2) and clinical data (column 4) for each phenotype. The odds path for very strong evidence, O_PVst_, associated with each prior, is reported in adjacent columns.

| Phenotype | Prior<br>(Population) | O<sub>PVst</sub><br>(Population) | Prior<br>(Clinical) | O<sub>PVst</sub><br>(Clinical) |
| --- | --- | --- | --- | --- |
| Familial hypercholesterolemia<br>(predicted using adjusted LDL ≥ 190) | 10.6% | 324 | 3.7% | 1444 |
| Breast or ovarian cancer | 9.8% | 363 | 6.8% | 616 |
| Colorectal cancer | 2.5% | 2483 | 5.6% | 814 |
| Hypertrophic cardiomyopathy | 0.2% | 64589 | 2.6% | 2397 |
| Dilated cardiomyopathy | 0.3% | 41301 | 0.7% | 15079 |
| Arrhythmogenic right ventricular<br>cardiomyopathy | 0.4% | 34238 | 1.1% | 7152 |
| Brugada syndrome | 0.2% | 74660 | 2.0% | 3342 |
| Li-Fraumeni syndrome<br>(predicted using presence of any cancer) | 23.2% | 103 | 5.6% | 813 |
| Maturity-onset diabetes of the young<br>(predicted using HbA1c ≥ 6.5%) | 7.0% | 585 | 7.2% | 565 |
| Long QT syndrome<br>(predicted using HbA1c ≥ 460) | 9.1% | 401 | 5.6% | 802 |
| Osler hemorrhagic telangiectasia syndrome | 0.1% | 195638 | 2.7% | 2271 |
| Juvenile polyposis syndrome<br>(predicted using presence of colon polyp) | 5.6% | 816 | 0.4% | 30907 |
| Pancreatic cancer | 0.3% | 48441 | 0.0% | inf |
| Prostate cancer | 3.0% | 1909 | 8.3% | 462 |
| Hereditary hemochromatosis | 0.2% | 81510 | 2.5% | 2508 |
| Ehlers-danlos syndrome | 0.2% | 61579 | 1.4% | 5305 |
| Retinitis pigmentosa | 0.1% | 237321 | 8.6% | 440 |
| Nephroblastoma | 0.0% | 2352837 | 1.0% | 8981 |

**Supplementary Table 3:** Prior probabilities of pathogenicity for aggregations. Prior probability of pathogenicity estimated based on population data (column 2) and clinical data (column 4) for for three aggregations across multiple phenotypes: across phenotypes with fewer than 25 pathogenic or benign ClinVar variants, across phenotypes associated with CDC tier one genes, and across all phenotypes. The odds path for very strong evidence, O_PVst_, associated with each prior, is reported in adjacent columns.

| Phenotype | Prior<br>(Population) | O<sub>PVst</sub><br>(Population) | Prior<br>(Clinical) | O<sub>PVst</sub><br>(Clinical) |
| --- | --- | --- | --- | --- |
| Phenotypes with fewer than 25 pathogenic or benign ClinVar variants | 1.4% | 5651 | 3.0% | 1962 |
| Phenotypes associated with CDC tier one genes | 8.6% | 438 | 5.4% | 852 |
| All phenotypes | 2.8% | 2120 | 1.9% | 3531 |

**Supplementary Table 4:**
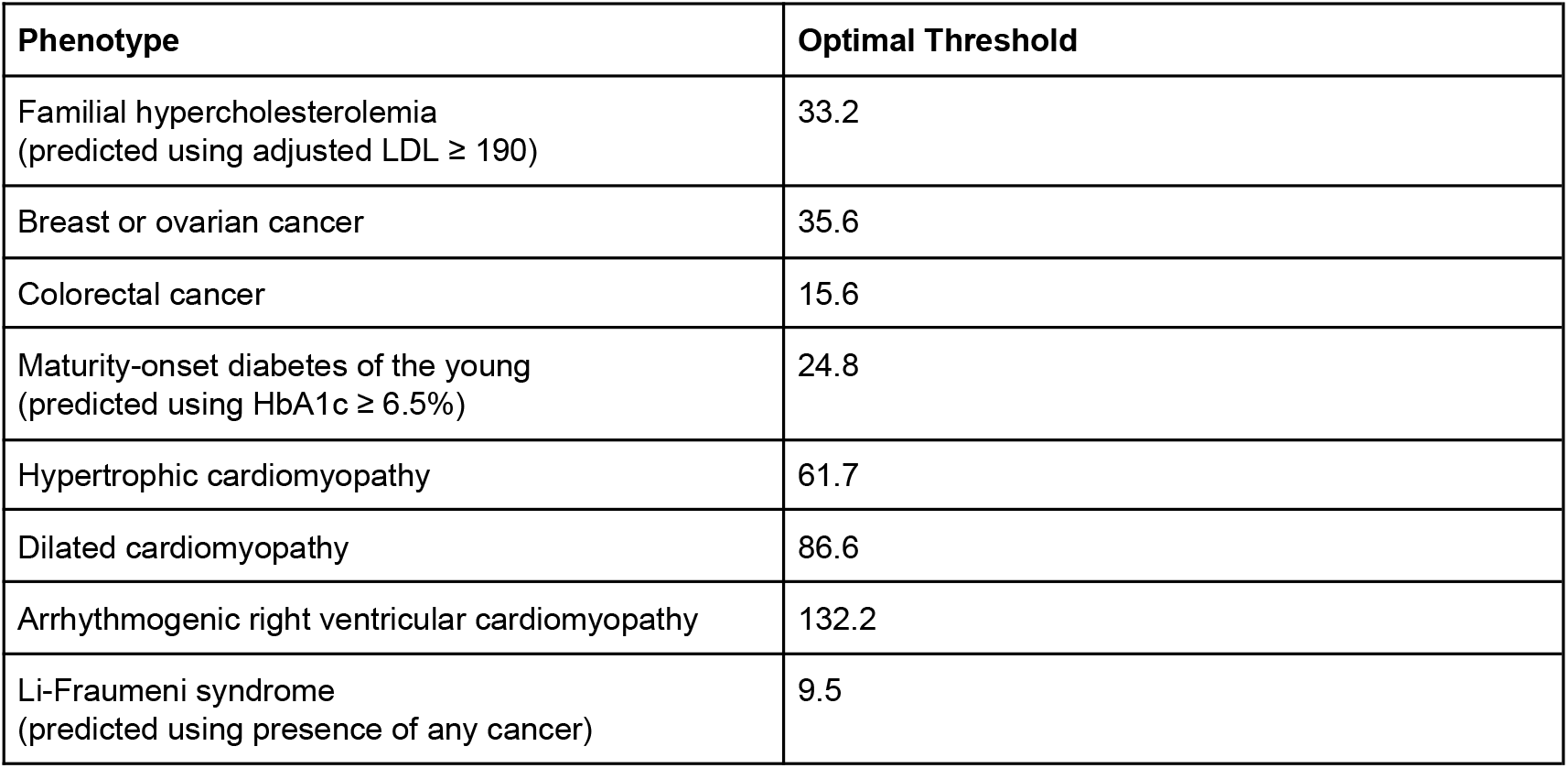
Optimal odds ratio thresholds for classification. Odds ratio thresholds at which specificity and sensitivity are maximized (using the Youden Index or most upper left point on each ROC curve, which can also be interpreted as the point at which global LR+ is maximized) for 8 phenotypes with at least 25 pathogenic or benign ClinVar variants.

| Phenotype | Optimal Threshold |
| --- | --- |
| Familial hypercholesterolemia<br>(predicted using adjusted LDL ≥ 190) | 33.2 |
| Breast or ovarian cancer | 35.6 |
| Colorectal cancer | 15.6 |
| Maturity-onset diabetes of the young<br>(predicted using HbA1c ≥ 6.5%) | 24.8 |
| Hypertrophic cardiomyopathy | 61.7 |
| Dilated cardiomyopathy | 86.6 |
| Arrhythmogenic right ventricular cardiomyopathy | 132.2 |
| Li-Fraumeni syndrome<br>(predicted using presence of any cancer) | 9.5 |

**Supplementary Table 5:** Estimated odds ratio evidence intervals at the gene level. We calculated odds ratio intervals that correspond to different ACMG/AMP evidence strength levels for 9 genes that had at least 25 pathogenic or benign ClinVar variants. A dash indicates that the specified level of evidence was not reached. N represents the number of ClinVar pathogenic and benign variants considered for calibration in each phenotype.

| Gene | N | Supporting | Moderate | Strong | Very Strong |
| --- | --- | --- | --- | --- | --- |
| LDLR | 94 | [6.1, 6.2] | [6.2, 6.6] | [6.6, 7.0] | $\geq 7.0$ |
| APOB | 52 | [-1.0, 0.2] | $\geq 0.2$ | – | – |
| BRCA1 | 115 | [4.9, 7.1] | [7.1, 23.2] | $\geq 23.2$ | – |
| BRCA2 | 143 | [5.0, 5.9] | [5.9, 9.5] | $\geq 9.5$ | – |
| ATM | 50 | [1.9, 1.9] | [1.9, 2.0] | [2.0, 2.1] | $\geq 2.1$ |
| MSH6 | 34 | [2.4, 2.8] | [2.8, 4.7] | [4.7, 6.6] | $\geq 6.6$ |
| MLH1 | 35 | [13.1, 20.5] | [20.5, 24.2] | [24.2, 28.0] | $\geq 28.0$ |
| TTN | 136 | [29.3, 57.4] | $\geq 57.4$ | – | – |
| TP53 | 59 | [2.6, 3.4] | [3.4, 6.2] | $\geq 6.2$ | – |

**Supplementary Figure 1:**
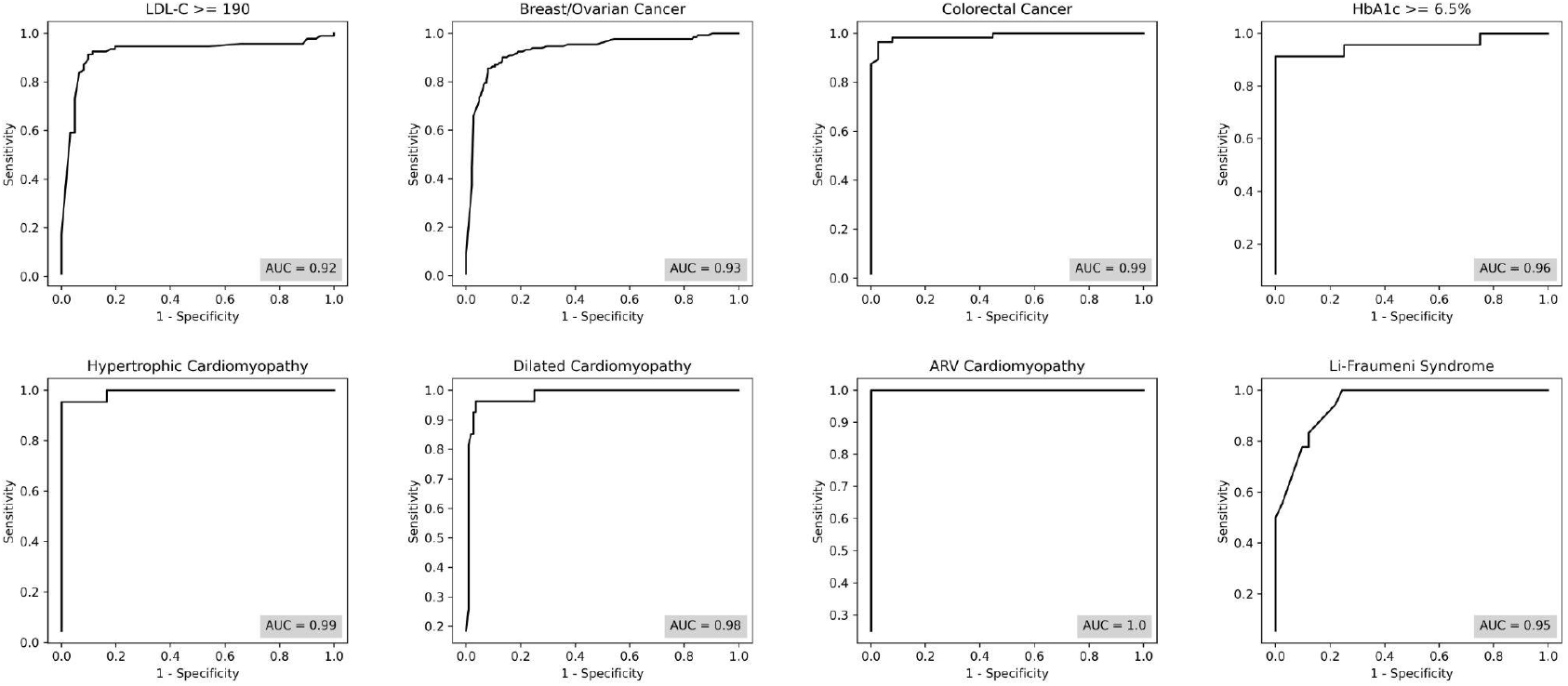
Variant classification ROC curves at the phenotypic level. ROC curves for classifying ClinVar variants using population odds ratios for 8 phenotypes with at least 25 pathogenic and benign variants. AUC ranged between 0.92 and 1.00 among these phenotypes.

**Supplementary Figure 2:**
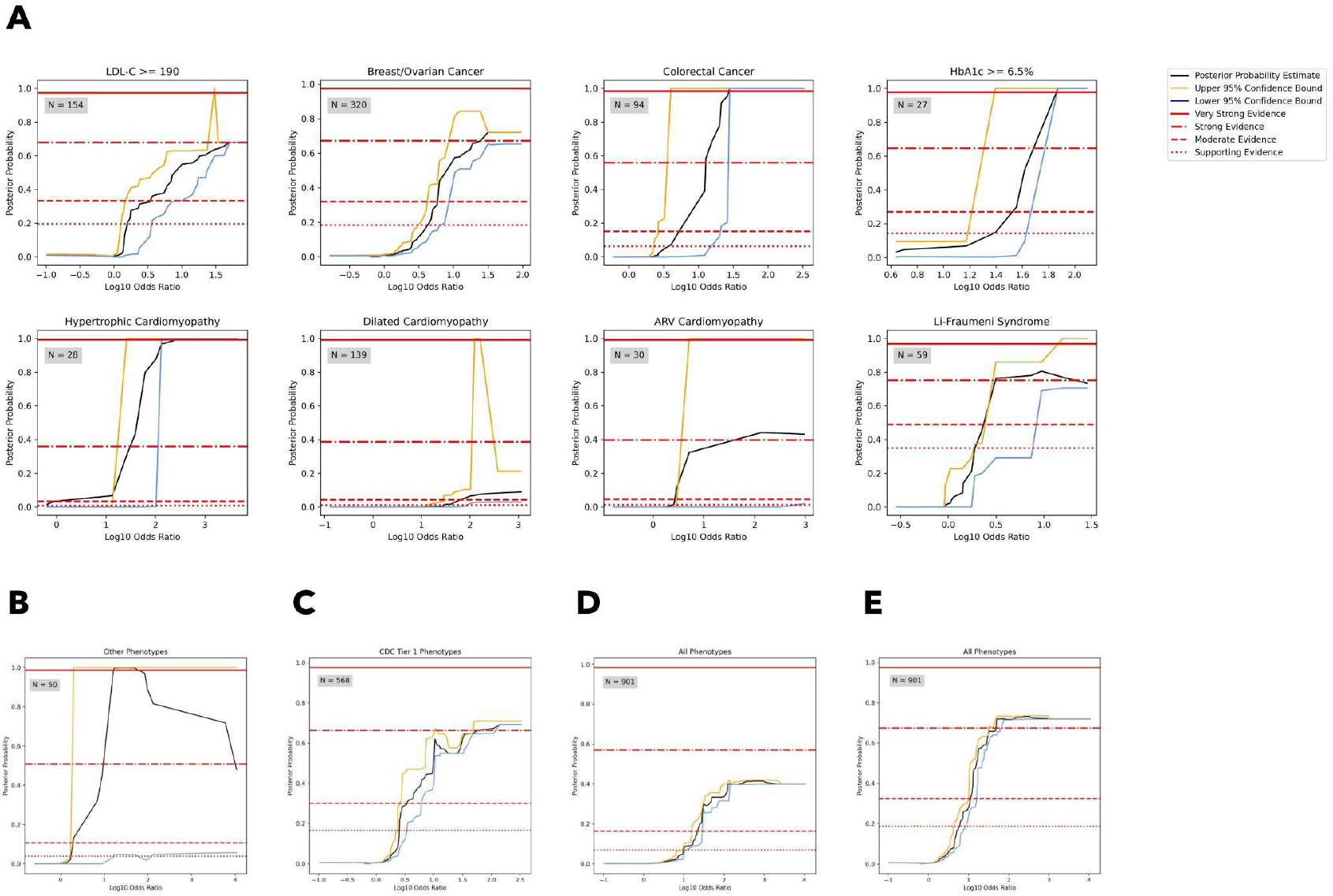
Bootstrapped posterior probability curves with 95% confidence bounds for selected phenotypes and aggregations. **[A]** Bootstrapped posterior probability curves for variants in 8 phenotypes with at least 25 pathogenic or benign ClinVar variants available for evidence calibration. **[B]** Bootstrapped posterior probability curve for variants in phenotypes with fewer than 25 pathogenic or benign ClinVar variants, in aggregate. **[C]** Bootstrapped posterior probability curves for variants in phenotypes associated with CDC Tier 1 genes, in aggregate. **[D]** Bootstrapped posterior probability curves for variants in all phenotypes, in aggregate. **[E]** Bootstrapped posterior probability curves for variants in all phenotypes, in aggregate, using a prior probability of pathogenicity of 10%. Three SD Gaussian smoothing was applied to all plots uniformly. Threshold levels used (supporting, moderate, strong, and very strong, from bottom to top) vary by plot based on the estimation of distinct priors for each phenotype and aggregation, as described in **Methods**. Posterior probability estimates and confidence bounds were derived via 10,000 bootstrapping iterations, where the posterior probability estimate, lower 95% confidence bound, and upper 95% confidence bound were the median, 5th percentile, and 95th percentile of the bootstrapped distribution, respectively.

**Supplementary Figure 3:**
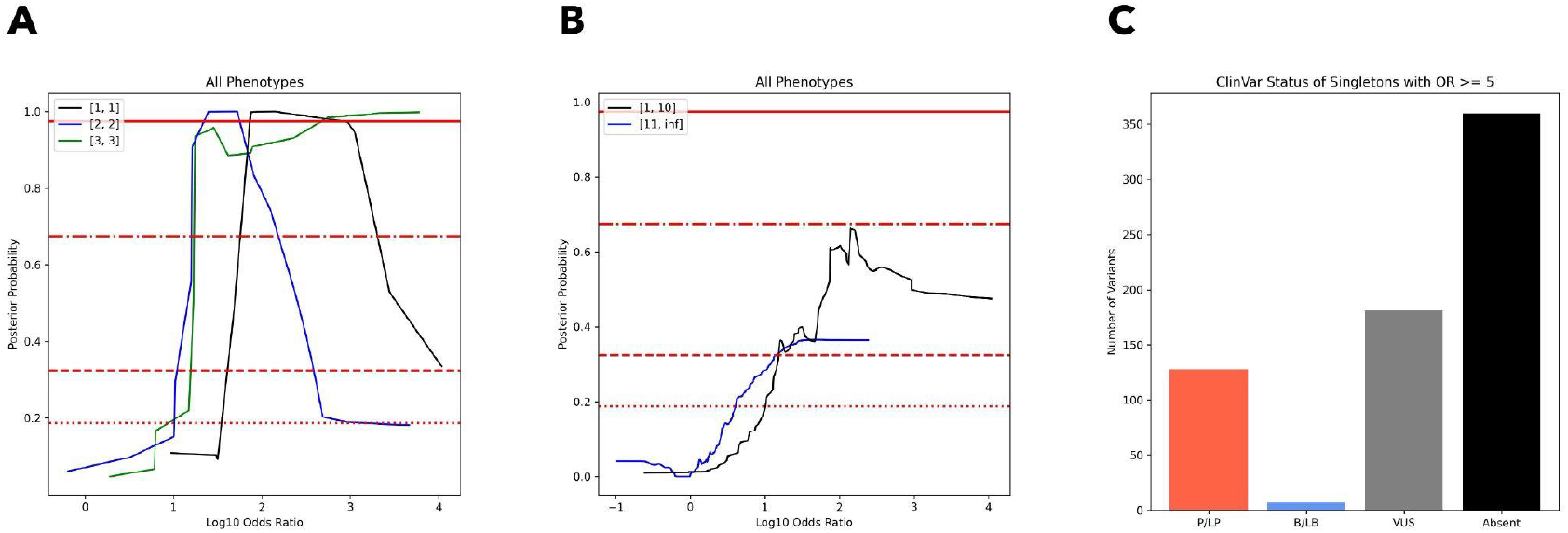
Singletons and variants with few patients can provide substantial evidence of pathogenicity. **[A]** Posterior probability curves for singletons, variants with two participants, and variants with three participants. **[B]** Posterior probability curves for variants with 10 or fewer participants and variants with more than 10 participants. Note that both classes of variants reach only moderate evidence, despite reaching strong evidence as a whole, because a larger dataset can benefit from shorter windows and a more equal distribution of pathogenic and benign variants (see **Methods** for details on calibration). **[C]** Distribution of OR ≥ 5 singletons by ClinVar status. All calibration was done using a prior of 10% and used the entire dataset across all phenotypes. Three SD Gaussian smoothing was applied to all calibration plots uniformly. Threshold levels used (supporting, moderate, strong, and very strong, from bottom to top) were based on the same 10% prior.

**Supplementary Figure 4:**
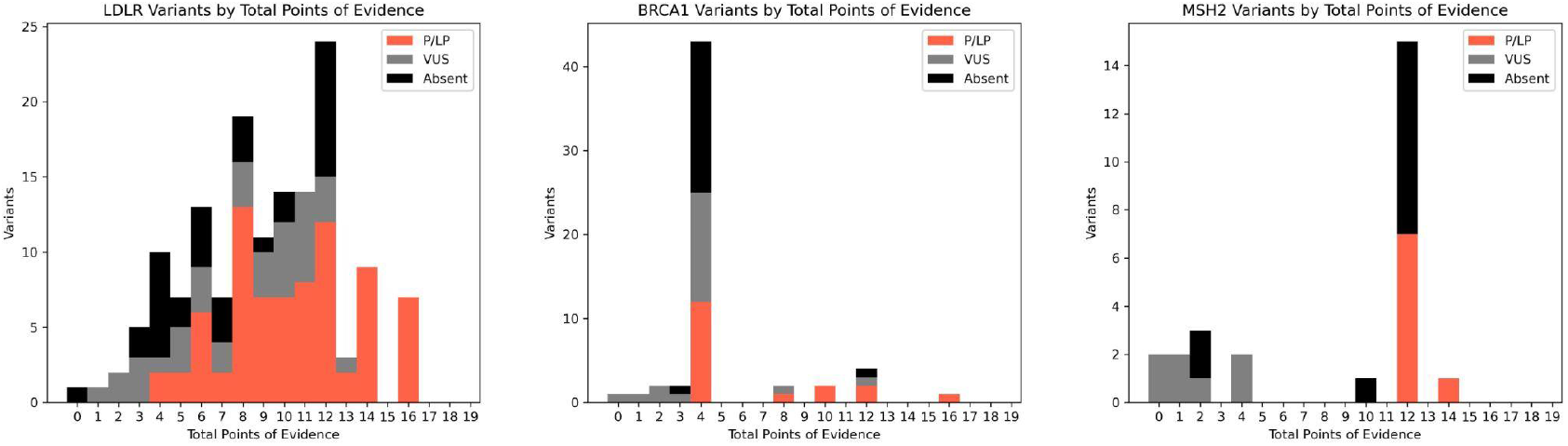
Number of points of evidence for select variants in *LDLR, BRCA1*, and *MSH2*. Applying the ACMG/AMP SVI guidelines, as described in **Methods**, we calculated the number of points of evidence from population, computational, functional, and contextual data sources for variants in *LDLR, BRCA1*, and *MSH2* that have ‘strong’ or ‘very strong’ population evidence, separated by ClinVar status. Notably, 60 VUS and variants absent from ClinVar reach ≥ 6 points, most of which are concentrated in *LDLR*.

## Notes

### Competing Interest Statement

The authors have declared no competing interest.

### Funding Statement

We gratefully acknowledge funding from NIH R01HG010372 (V.B., T.Y., L.B., C.A.C.), NIH R01HG013350 (V.P.), NIH R56HG012681 (T.Y., C.C.), and AHA 24TPA1300072 (T.Y., C.C.).

### Author Declarations

Mass General Brigham IRB gave ethical approval for this work (protocol 2020P002093). Work was performed under UK Biobank application 41250.

### Summary of Updates

We have made substantial revisions and introduced important new analyses. We initially focused on three CDC 'Tier 1' conditions, but have now expanded to a broad set of 18 clinically actionable phenotypes covering 41 genes from the ACMG Secondary Findings v3.2 list. This significantly broadens the clinical relevance of our findings. We also calibrated phenotype-level evidence thresholds for odds ratios, enabling precise application of population for clinical interpretation. We also performed detailed analyses on gene-level evidence for rare variants within specific phenotypes. Additionally, to address concerns regarding increased variance when using small allele counts, we conducted sensitivity analyses evaluating the robustness of our evidence thresholds for ultra-rare variants.

